# Intrinsic Capacity of Rural Elderly in Thar Desert using WHO ICOPE (Integrated Care for Older Persons) Screening Tool: A Pilot Study

**DOI:** 10.1101/2022.02.04.22270231

**Authors:** Arvind Mathur, Pankaj Bhardwaj, Nitin Kumar Joshi, Yogesh Kumar Jain, Kuldeep Singh

## Abstract

**Introduction:** ICOPE (Integrated Care for Older Persons) screening tool helps to address declines in physical and mental capacities in older people. In India majority of the older population resides in rural areas and there is a paucity of studies that demonstrates the utility of the ICOPE screening tool in India. Thus, this study was conducted to demonstrate the feasibility of using WHO ICOPE screening tool in a rural population.

**Methods:** A cross-sectional study was conducted in the two villages of Jodhpur. WHO ICOPE screening was done for a comprehensive geriatric assessment of intrinsic capacity by determining cognitive decline, the decline in mobility, malnutrition, sensory loss, depressive symptoms, health risks, social care, and support. Additionally, demographic details and information about health conditions were gathered.

**Result:** A total 451 participants (Male=54.5% and Female= 45.5%) surveyed. The mean age was 68.36 (SD=7.73) years and the majority of participants (68.3%) were illiterate. Most of the elderly have good families and lived with their children. Cognitive decline was found to be prevalent in 31.5% (n=142) of the participants, while mobility was diminished in 52.1% (n=235) participants. Eye problems were observed in 49.4% (n=223) participants, while 68.3% (n=308) were observed to be suffering from hearing loss. Limitation in mobility observed with the chair rise test and feeling of pain was found to be significantly associated with gender difference, with more limitation in females than males (p = 0.005; χ2 = 7.95) and more pain felt by females than males (p = 0.001; χ2 = 15.64).

**Conclusion:** This tool seems suitable in identifying the intrinsic capacity of the rural elderly.

## Introduction

Population aging is one of the most concerning demographic phenomena of the current times with an ever increase in geriatric population as a result of declining fertility and increasing longevity.^1^ Reports from the World Health Organization project 1 amongst every 6 people to be aged 60 or above by the time the world witnesses 2030, which again is expected to double by 2050.^2^ This phenomenon is of concern as it comes associated with its own set of public health issues related to health conditions, diminishing intrinsic capacity, care and finance related hardships.^2,3^

The situation presents an even greater challenge in middle income countries like India, where the elderly population has risen by 32.7% in the last decade in contrast to the rise of general population by 12.4%. This corresponds to an increase of a staggering 34 million people aged 60 years and above. Moreover, MOSPI projects the number to increase by another 56 million over the coming decade.^4^

WHO defines healthy ageing as having a functional ability to do what a person values. Such an ability is essential for one’s well-being and the its core drivers are the intrinsic capacity of the individual and the interaction with the environment around them.^5^ In continuation with the above definition, the ICOPE approach was recommended by the WHO (2017), emphasising the focus on improving intrinsic capacity and functional ability of an individual as the key towards healthy ageing. The recommendations could further serve as the basis for support and inclusion in the national guidelines, primary care programmes and essential care packages for universal health coverage to prevent care-dependency.^6^

ICOPE (Integrated Care for Older Persons) screening tool helps to address declines in physical and mental capacities in older people. In India majority of older population resides in the rural areas and there is a paucity of studies which demonstrates the utility of ICOPE screening tool in India. Thus, this study was conducted to demonstrate the feasibility of the community-based screening of older people using WHO ICOPE screening tool in a rural Indian population.

## Methodology

A cross sectional study was conducted in the Jodhpur, which is also called as gateway of Thar Dessert and is amongst the highest populated districts of Rajasthan – the largest state of India. Two rural villages selected from different tehsils of Jodhpur. After due approval from the Institutional Ethical Committee of All India Institute of Medical Sciences, Jodhpur (Rajasthan, India; IEC Certificate Number: AIIMS/IEC/2020-21/3075), a total of 451 geriatric people aged 60 years and above were included in the study through a home-based survey. After due consent, WHO ICOPE screening was done for a comprehensive geriatric assessment of intrinsic capacity by determining cognitive decline, decline in mobility, malnutrition, sensory loss, depressive symptoms, health risks, social care and support. Additionally, demographic details and information about any habits were gathered through interviews and physical examination was carried out to measure vitals – blood pressure, random blood sugar and ulnar height. Impact on caregivers was recorded through a questionnaire and risk of abuse of elderly was recorded through careful observation by the interviewer.

As per the WHO ICOPE Screening tool, cognitive decline was determined if there was difficulty or failure in answering to questions related to orientation of time and place or recalling words; decline in mobility was assessed through the chair rise test; and malnutrition was assessed through recent weight loss or loss of appetite. Sensory loss was determined through the assessment of visual and hearing impairment and depressive symptoms were assessed through scoring patients’ perception of feeling down, depressed, hopeless and lack of interest in doing things over past 2 weeks. The components of the intrinsic capacity were scored either 0 – representing decline or 1 – representing no decline.

The data was presented as mean, standard deviation, numbers and percentages. Difference in characteristics was evaluated using chi-square test for categorical and t-test for continuous variables. The statistical analysis was performed using SPSS (version 23) and the p-value less than 0.05 was considered significant.

## Results

A total of 451 participants were included in the study and were found to have a mean age of 68.36 (SD=7.73) years ranging from 60 years to 98 years. Amongst these, males were 54.5% (n=246) with the mean age of 67.84 (SD: 6.68; range: 60-98) years and females were 45.5% (n=205) with the mean age of 68.98 (SD: 8.81; range: 60-98) years.

Maximum number of participants (n=175; 38%) were aged between 60-64 years. Out of all participants surveyed, 76.1% (n=343) were married and 23.1% (n=104) were widowed, while 0.9% (n=4) participants were single. Majority of the participants (68.3%; n=308) were illiterate and earned their livelihood doing farming (40.8%; n=184). Almost every participant (91.4%; n=412) lived with children and their family and did not face any problem related to accommodation (94.9%; n=428), nevertheless, 12.2% (n=55) faced difficulties due to finance related issues. The demographic details of the study participants is shown in table 1.

**Table 1:** Demographic Details of the Study Participants.

|  | Frequency (n) | Percentage (%) |
| --- | --- | --- |
| **Gender** |  |  |
| Males | 246 | 54.5 |
| Females | 205 | 45.5 |
| **Age Distribution** |  |  |
| 60-64 | 175 | 38.8 |
| 65-69 | 90 | 19.96 |
| 70-74 | 90 | 19.96 |
| 75-79 | 41 | 9.1 |
| 80-84 | 38 | 8.4 |
| 85-89 | 8 | 1.8 |
| 90-94 | 7 | 1.6 |
| 95-99 | 2 | 0.4 |
| **Marital Status** |  |  |
| Single | 4 | 0.9 |
| Married | 343 | 76.1 |
| Widowed | 104 | 23.1 |
| **Education** |  |  |
| Illiterate | 308 | 68.3 |
| Literate (no Formal Education) | 38 | 8.4 |
| Primary Education | 77 | 17.1 |
| Secondary Education | 21 | 4.7 |
| Graduation | 5 | 1.1 |
| Post Graduate | 2 | 0.4 |
| **Living Status** |  |  |
| Alone | 8 | 1.8 |
| With Spouse Only | 26 | 5.8 |
| With Children and Family | 412 | 91.4 |
| Old Age Home | 5 | 1.1 |
| **Occupation** |  |  |
| Farmer | 184 | 40.8 |
| Unemployed | 106 | 23.5 |
| Housewife | 104 | 23.1 |
| Self Employed | 37 | 8.2 |
| Labourer | 9 | 2.0 |
| Retired | 6 | 1.3 |
| AWW | 2 | .4 |
| Government Employee | 2 | .4 |
| Sarpanch | 1 | .2 |

Upon assessment of the intrinsic capacity, cognitive decline was found to be prevalent in 31.5% (n=142) of the participants, while mobility was diminished in 52.1% (n=235) participants. Eye problems was observed in 49.4% (n=223) participants, while 68.3% (n=308) were observed to be suffering from hearing loss. Recent weight loss was reported by 17.5% (n=79) individuals while appetite loss was reported by 33.7% (n=152) individuals. Though the self-reported health was “Good” by 39.7% (n=179) individuals and “Fair” by 38.4% (n=173), nearly half 46.8% (n=211) of the participants experience symptoms of some illness, with 63.0% (n=284) having a pre-existing systemic disease. Pain was experienced by 50.8% (n=229) of the respondents with a history of fall amongst 9.5% (n=43) respondents in the past 12 months. Inability to sleep well was reported in 21.3% (n=96) and 19.3% (n=87) reported feeling depressed. Despite these values, 88.5% were able to undertake daily activities independently and 98.9% (n=446) caregivers did not feel that the elderly had a negative impact on their lives. Intrinsic capacity decline is illustrated in table 2.

**Table 2:**
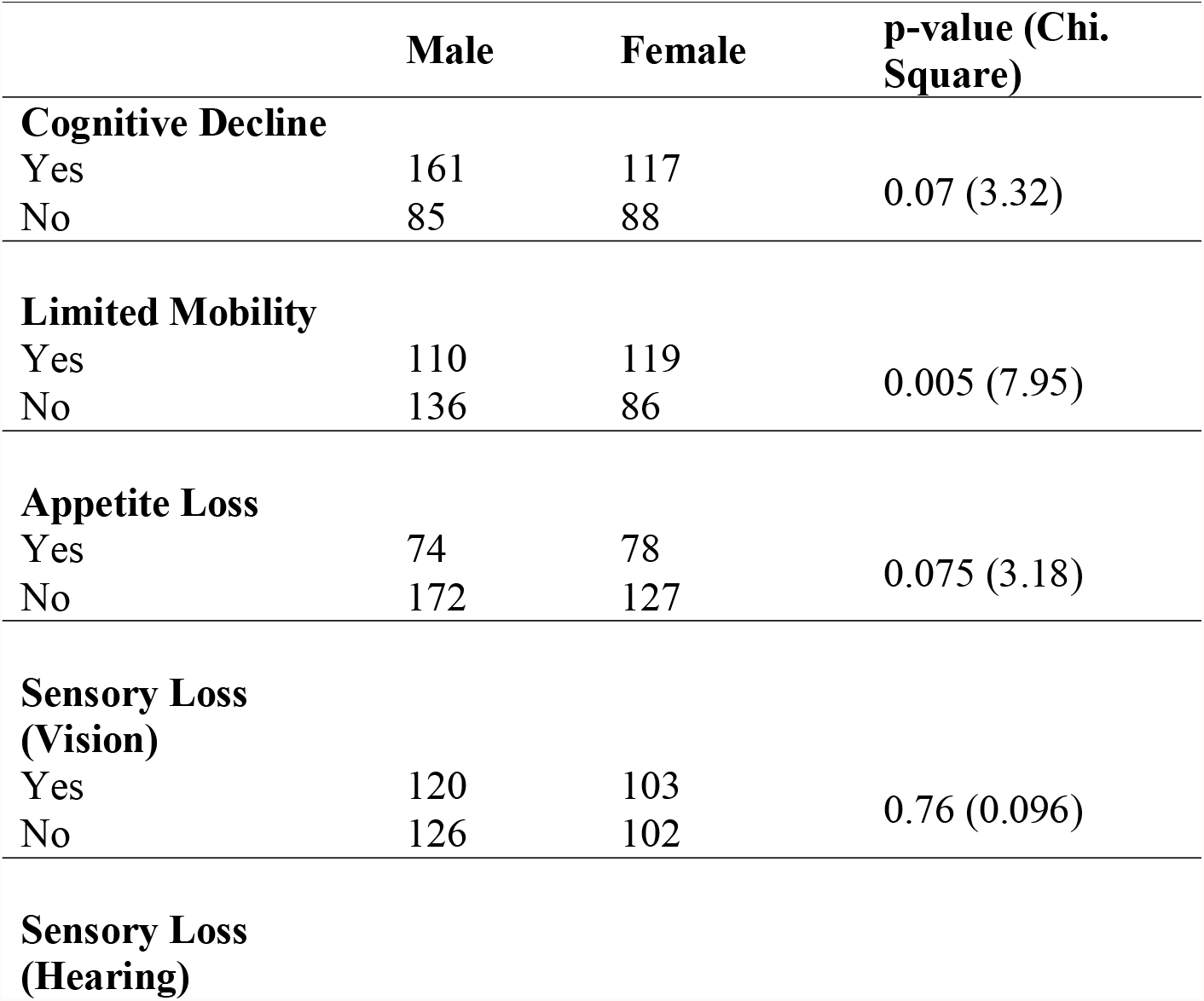
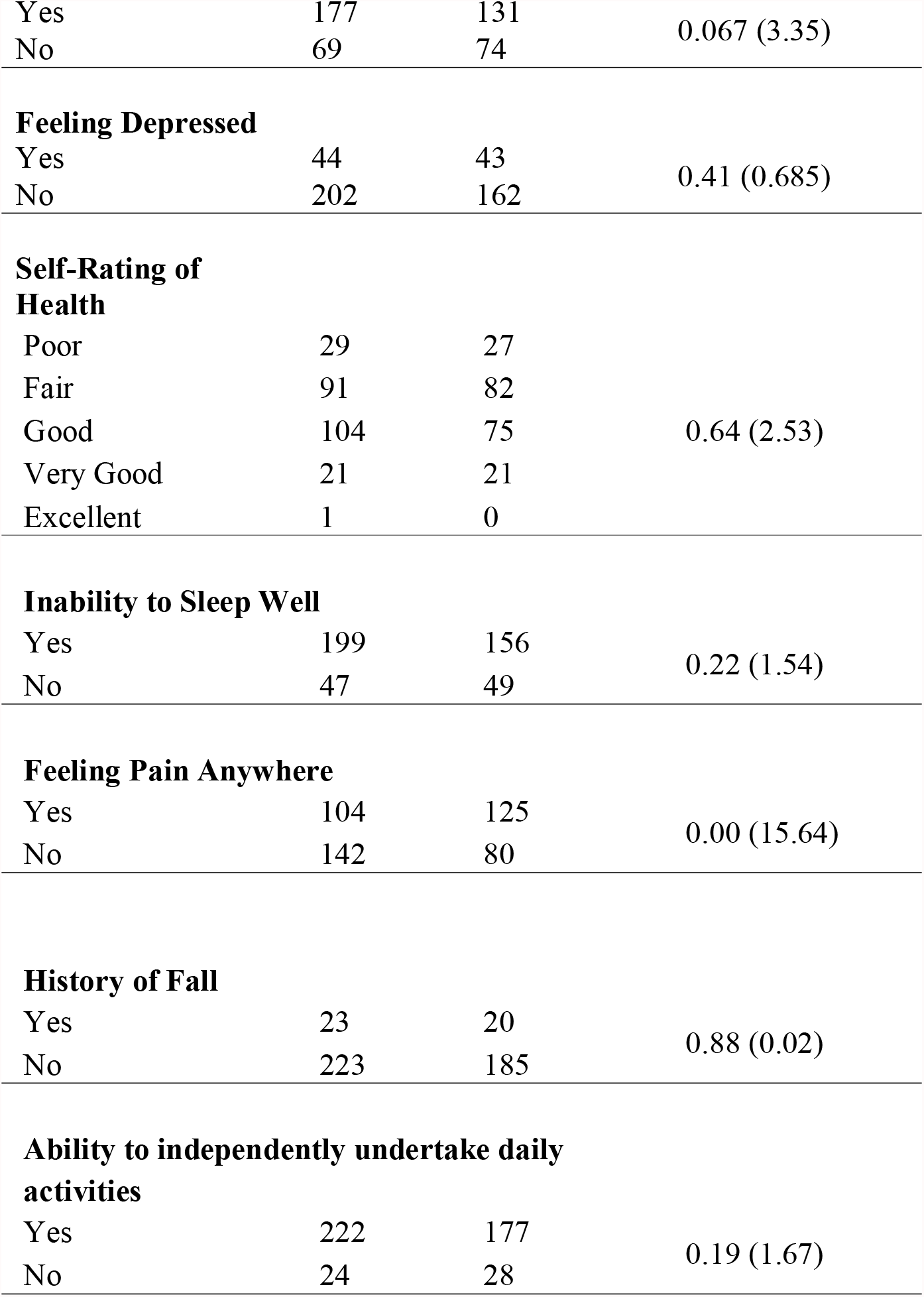
Intrinsic Capacity Decline as per the ICOPE Screening Tool.

Limitation in mobility observed with the chair rise test and feeling of pain were found to be significantly associate with gender difference, with more limitation in females than males (p = 0.005; χ^2^ = 7.95) and more pain felt by females than males (p = 0.001; χ^2^ = 15.64).

More than half (n=321; 71.2%) of the surveyed population were found to be using tobacco products of some kind (smokeless, smoked or both, while, 12 (2.7%) participants reported to consume alcohol daily.

A quarter of the study participants (n=124; 27.5%) were found to have an elevated systolic blood pressure (>140 mm of Hg) suggestive of systolic hypertension, out of whom, 66 (32.2%) were females and 58 (23.6%) were males. Elevated blood sugar levels were observed in 23 (5.1%) participants with 8 (3.9%) females and 15 males (6.1%) as shown in table 3.

**Table 3:**
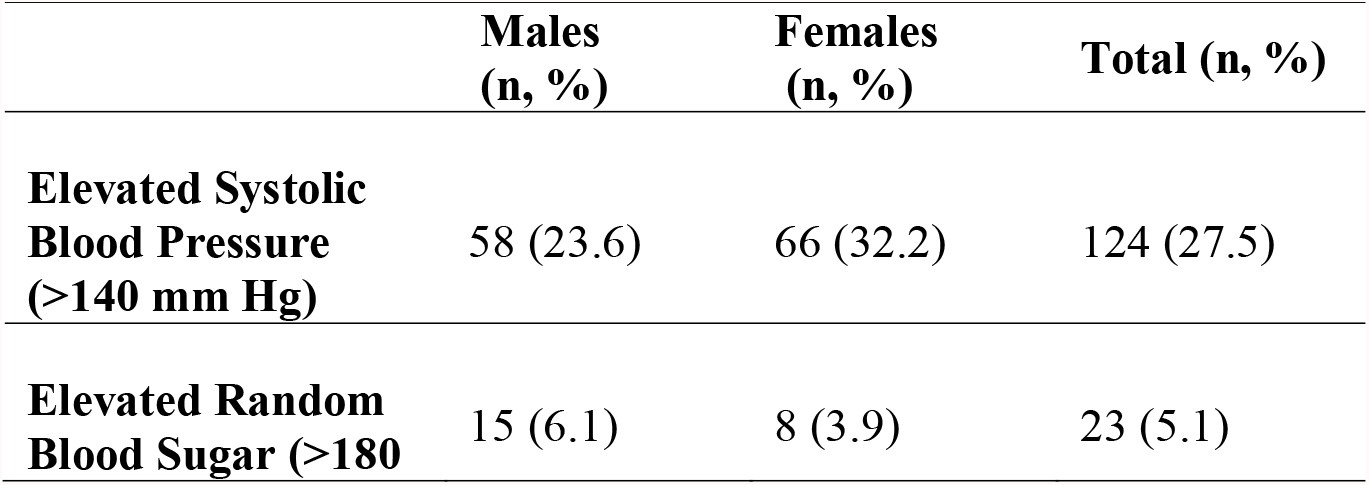
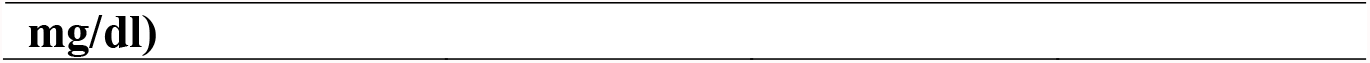
Blood Pressure and Random Blood Sugar of Study Participants.

More than a third study participants (n=180; 40%) did not require caregivers support while another third (n=165; 36%) admitted that they were not able to undertake any domestic activities like cooking and shopping. More than 90% of respondents were observed to have no risk of abuse and had a supportive caring family.

## Discussion

This feasibility study revealed that majority of the participants were illiterate and earned their livelihood doing farming. These results are line with others surveys and studies conducted among rural elderly population of India.^7,8.^ Various studies have mentioned that in elderly population in rural India is far more illiterate as compared to elderly living in urban areas.^7,8^The low literacy status negatively impacts the healthcare utilisation and make rural elderly more vulnerable in terms of having poor health outcomes.^7,9,10^ Results of this study also coincides with the findings of the other studies that mentions that most elderly in rural areas are mostly self-employed in the agriculture sector and related occupations and have with no retirement age or pension benefits.^7,8,11^ Poor and illiterate elderly in rural areas have a higher rate of untreated morbidities due to ack of access and means to use health care facilities.^7,10^ ICOPE screening tool can be very useful for identifying poor physical and mental function in such population experience chronic conditions.^12^ It was also evident from the study that rural elderly has good family support as most of them lived with their children and do not have accommodation related issue. Studies suggest that aged who live with the children have good health status and lower incidence of ailment as compared to those who do not live with their children.^7,13,14^ Though families of the elderly are envisaged to provide the necessary care and assistance for the elderly but detailed study of recent changes in the structure of families and reorganisation of the duties and functions of family members is needed to gauge the scenario.

In this study cognitive decline was found in more than thirty percent of the rural elderly which is considerably higher than depicted in other studies conducted in other states of India. The reason for this might be attributed to the method used is determining Cognitive decline.^15^ In ICOPE study cognitive decline is determine if participants provided a wrong response to either of the two questions on orientation in time and space or if they could not recall the three words they were asked to remember whereas in other studies cognitive decline were assessed using MMSE scale.

No significant difference was seen in gender and cognitive decline. There has been few research in India on gender differences in cognitive health among older persons, and the results have been equivocal.^16^

More than half of the participants reported to have limited mobility. Though there is paucity of studies about limited mobility in elderly in India but evidences from cross-sectional studies on fall among elderly reported to have annual fall in more than half of the surveyed population. Studies have shown that mobility impairment is associated increased risk of falls.^17,18^ As there is lack of data on limited mobility from rural populations, more studies are need.

Eye problems was observed in half of the participants. Similar finding were disclosed in the first Longitudinal Ageing Study in India.^19^ This study provided insight into the extent of vision-related disorders among India’s elderly. According to LASI study 45% of elderly has vision related problems in state of Rajasthan. In the elderly population, non-working status, economic dependency, illiteracy, and tobacco use are supposed be to linked to visual impairment. Those with visual impairment have a lower quality of life when it comes to eyesight. Studies have shown that, elderly people with eye problems have lower quality of life and are more likely to be depressed.^20^ There is evidence that having poor vision increases the risk of falling in elderly people.^21^

According to the WHO, older persons and people of all ages with pre-existing medical issues are more likely than others to suffer serious illness. This study revealed that more than sixty percent elderly has pre-existing systemic disease. India Report on Longitudinal Ageing Study of India (LASI) Wave-1 also revealed that more than 70% elderly in India suffer from chronic diseases.^19^ By 2030, the elderly are predicted to bear 45 percent of the entire burden of diseases, the majority of which are non-communicable. Therefore, adequate investment in aged healthcare and effective policies, as well as their timely management, are critical.

## Conclusion

The WHO ICOPE screening tool have provided a good yield in identifying intrinsic capacity of rural elderly. This tool seems suitable in identifying rural individuals with poor physical and mental function.

## Data Availability

All data produced in the present study are available upon reasonable request to the authors

